# Risk Stratification for Ventricular Tachyarrhythmia in Patients with Non-Ischemic Cardiomyopathy

**DOI:** 10.1101/2023.08.30.23294871

**Authors:** Ido Goldenberg, Arwa Younis, David T. Huang, Spencer Rosero, Valentina Kutyifa, Scott McNitt, Bronislava Polonsky, Jonathan S. Steinberg, Wojciech Zareba, Ilan Goldenberg, Mehmet K. Aktas

## Abstract

**Background:** The implantable cardioverter defibrillator reduces mortality among patients with heart failure (HF) due to ischemic heart disease. Clinical trial data have called into question the benefit of an ICD in patients with HF due to non-ischemic cardiomyopathy (NICM).

**Objective:** We developed a risk stratification score for ventricular tachyarrhythmia (VTA) among patients with NICM receiving a primary prevention ICD.

**Methods:** The study population comprised of 1,515 patients with NICM who were enrolled in the landmark MADIT trials. Fine and Gray analysis was used to develop a model to predict the occurrence of VTAs and ICD therapies while accounting for the competing risk of non-arrhythmic mortality. External validation was carried out in the RAID Trial population.

**Results:** Four risk factors associated with increased risk for VTA were identified: male sex, left ventricular ejection fraction ≤25%, no indication for cardiac resynchronization therapy with a defibrillator (CRT-D), and Black race. A score was generated based on this model and patients were stratified into low (N=390), intermediate (N=728), and high-risk (N=387) groups. The five-year cumulative incidences of VTA were 15%, 24%, 42% respectively. Application of score groups for the secondary endpoints of Fast VT or VF and Appropriate ICD Shock revealed similar findings. Recurrent event analysis yielded consistent results. The AUC in the validation cohort for the endpoint of Appropriate ICD Shock was 69.3.

**Conclusions:** Our study shows that patients with NICM can be risk stratified using demographic and clinical variables and may be used when evaluating such patients for a primary prevention ICD.

**CONDENSED ABSTRACT:** Data regarding the effectiveness of the ICD is lacking in patients with HF and NICM. The purpose of this study was to develop a risk score for ventricular tachyarrhythmia (VTA) among patients with NICM. We included patients from the MADIT trials to generate a risk score. Four risk factors associated with increased risk for VTA were identified and incorporated into the score. Patients were stratified into low, intermediate, and high-risk groups. The five-year cumulative incidences of VTA were 15%, 24%, 42% respectively. Our risk prediction model can support patient-physician shared decision making regarding primary ICD implantation in patients with NICM.

## INTRODUCTION

The benefit of the implantable cardioverter defibrillator (ICD) in patients with ischemic cardiomyopathy with reduced left ventricular ejection fraction (LVEF) has been widely studied and established(1). In contrast, for patients with non-ischemic cardiomyopathy (NICM) there is a paucity of randomized control trial data supporting its use(2). The Defibrillator in Patients with Nonischemic Cardiomyopathy Systolic Heart Failure (DANISH) trial is the most recent randomized clinical trial to assess the survival benefit of the ICD in NICM patients. The study revealed no difference in all cause-mortality between ICD recipients and non-ICD recipients(3). These recent data led to differences in guideline recommendation for primary ICD implantation in NICM patients, wherein the US guideline provide a Class I recommendation in patients with LVEF ≤ 35% with New York Heart Association (NYHA) Class II/III whereas European guidelines downgraded this recommendation to Class IIA(4).

The goal of our study was to develop and externally validate a risk stratification scheme for primary prevention in ICD implantation in patients with NICM, that may provide a simple clinical tool for shared decision making on the need for primary device implantation in this controversial population.

## METHODS

### Study Population

The model development cohort comprised 1515 patients with NICM implanted with a primary prevention ICD with or without cardiac resynchronization therapy (CRT) who were enrolled in the following 2 randomized, multicenter clinical trials (illustrated in Figure 1): Multicenter Automated Defibrillator Implantation Trial Cardiac Resynchronization Therapy(5) (MADIT-CRT), and Reduce Inappropriate Therapy(6), (MADIT-RIT). The studies were conducted between July 2009 through December 2011. The design and results of each of these trials have been reported previously. MADIT-CRT(5) enrolled 1814 patients with ischemic or NICM a LVEF ≤ 30%, a QRS duration of ≥130 milliseconds, and New York Heart Association (NYHA) functional class I or II symptoms. Patients were randomly assigned in a 3:2 ratio to receive CRT-D (1083 patients) or an ICD only (731 patients). MADIT-RIT(6) enrolled 1465 patients from 98 centers, and all patients met guideline criteria to receive an ICD or CRT-D for the primary prevention of sudden cardiac death. Only patients with NICM and those receiving a primary prevention ICD were included in this analysis. Each study was approved by the institutional review boards (IRB) at each enrolling site before participation. All patients provided informed consent before enrollment into the respective trials. A description of each trial is provided in Supplementary Table A.

**Figure 1:**
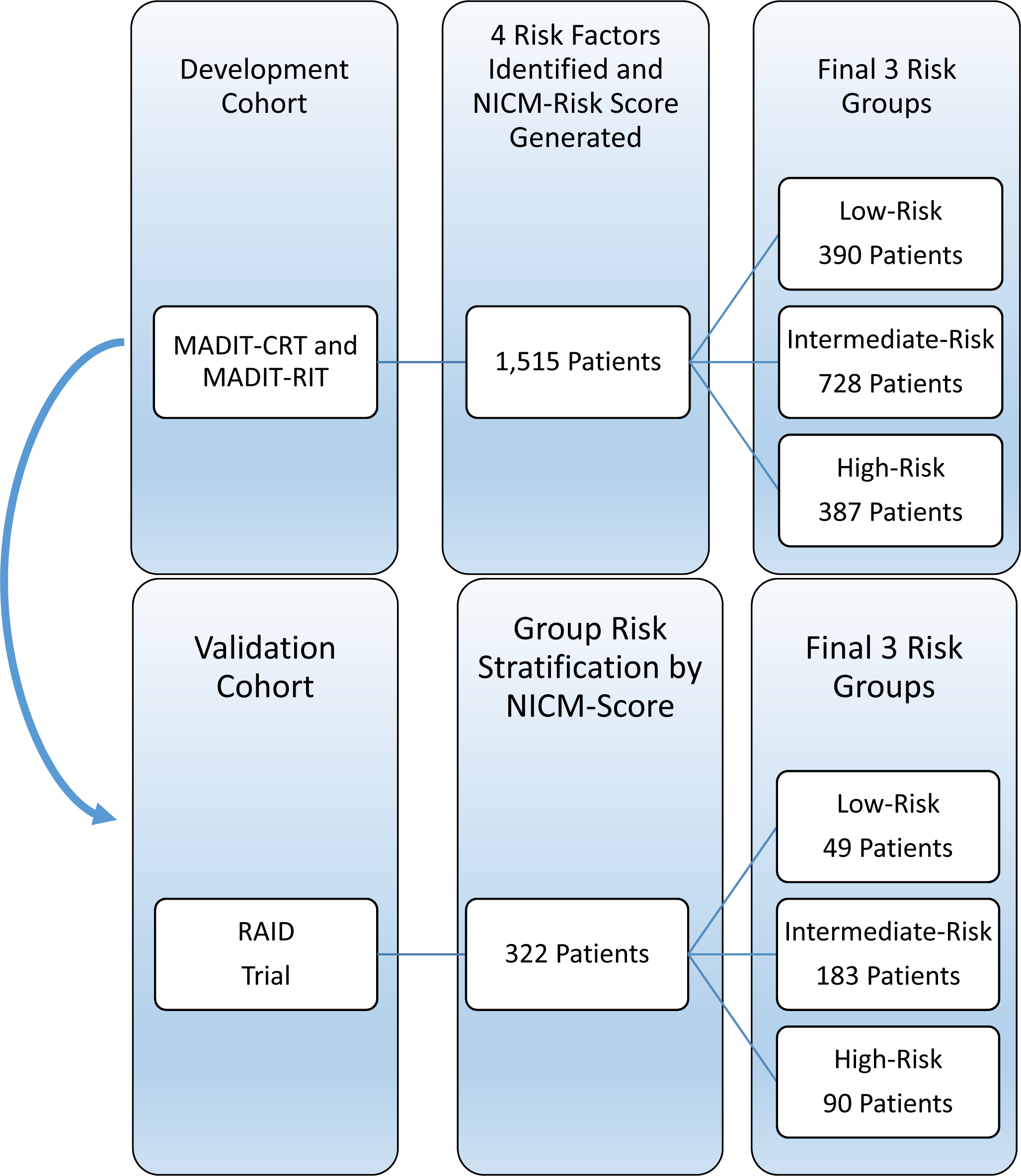
Study Population and Methodology.

External validation was carried out in 332 patients implanted for primary prevention in the Ranolazine in High-Risk ICD Patients (RAID^6^) trial conducted between 2011 and 2017.

### Endpoints and Arrhythmia Adjudication

All arrhythmia episodes and ICD therapies delivered in each of these trials were blindly adjudicated by at least two experienced electrophysiologists.

The primary endpoint for this study was any ventricular tachycardia (VT) or ventricular fibrillation (VF) defined as ICD recorded, treated (anti-tachycardia pacing and/or shock), or monitored sustained VT ≥170 beats/minute (bpm) or VF. Please see Supplementary Table B for adjudication protocol and definitions of all arrhythmic endpoints. Secondary endpoints included:

A. Fast VT/VF defined as VT≥200 bpm or VF.
B. Appropriate ICD Shock defined as ICD shock therapy for sustained VT≥200 bpm or VF.
C. The burden of arrhythmic endpoints, assessed in a recurrent events analysis.

Death without experiencing the arrhythmic endpoint was treated as the competing event in the analysis.

### Statistical Analysis

Continuous variables are expressed as mean+/-standard deviation. Categorical data are summarized as frequencies and percentages. Baseline characteristics are presented and compared by having experienced the primary endpoint of the study (Any VT/VF).

#### Predictive Model Development

We initially identified factors associated with increased risk for Any VT/VF in patients with NICM, using best subsets selection (p<0.05 was determined as sufficient to enter the final model Fine and Grey modeling that accounts for the competing risk of death). We then constructed a predictive model for the occurrence of Any VT/VF with these factors.

Numeric variables were made binary by the use of cut-off points with the goal of finding a simple, easily implemented scoring method to risk stratify patients. Thresholds for categorization of numeric variables were pre-specified using well-accepted clinical criteria.

After selection of binary covariates, they were used in a Fine and Gray model with VT≥170 bpm or VF (Any VT/VF) as the primary end point and death without Any VT/VF as the competing end point. Each covariate was then assigned a numeric value based on the relative value of its regression coefficient in the multivariate model. Specifically, each regression coefficient was multiplied by 10 to obtain the number of points associated with having the covariate. A VTA risk score was constructed by adding the assigned numeric values of the identified factors in each patient, and the study population was divided based on patient-specific score.

Based on the distribution of the score in our study population, we categorized each patient into either a low-, intermediate, or high-risk group, respectively. The score cutoffs for the groups were obtained as follows: 1) those below the 25^th^ percentile were sorted into the low-risk group, 2) those between the 25^th^ and 75^th^ percentile were sorted into the intermediate risk group, 3) and those with a score above the 75^th^ percentile were sorted into the high-risk group.

Cumulative incidence function curves (CIF) were used to estimate the risk of Any VT/VF in each of the 3 risk groups. The risk groups were further compared in a Fine and Gray regression model using hazard-ratios for the specific endpoints.

Finally, we carried out a recurrent event analysis where the burden of events in each group was displayed with a mean cumulative function (MCF) curve, estimating the mean number of events per person. Anderson-Gill modeling was used to generate hazard ratios to compare the burden of events between the 3 risk groups.

#### Calibration and Validation of the Model

##### A. Development Cohort

The internal validation of the development cohort was carried out by the k-fold cross-validation method, which has been also applied to competing risks regression models(7). We partitioned the data into 100 equal sized subsamples and of the 100 subsamples a single subsample was retained as the validation data for testing a model with the remaining 99 subsamples to be used as training data. The sampling size was set to 1000. This method was repeated using bootstrapping 100 times with each of the 100 subsamples serving as the validation data set exactly once. Finally, Brier score and area under the curve (AUC) were generated to quantify a prediction accuracy index(7). A calibration plot was also used to compare the predicted probability with the observed probability at a certain time point. The indices and plots are those estimated after 5 years of follow up and were generated for each of the study endpoints.

##### B. Validation Cohort

The external validation cohort comprised patients with implanted with a defibrillator with non-ischemic cardiomyopathy for primary prevention in the RAID trial. We generated calibration plots and quantified the predictive accuracy of the models using the Brier score and the AUC estimation at 2.8 years of follow up for all study endpoints. Importantly, the study did not show a significant effect of ranolazine on the risk of VT/VF and/or death.

All statistical tests were two sided, a p-value of <0.05 was considered statistically significant. Analyses were carried out with SAS software (version 9.4, SAS institute, NC, USA) and R Studio.

## RESULTS

### Study Population

Among 1515 patients with NICM who comprised the model development cohort, mean age was 59 ± 12 years, and 709 (38%) were women. Baseline clinical characteristics by those with and without VT/VF are presented in Table 1. Patients who experienced Any VT/VF were significantly more likely to be males and have a prior history of NSVT or atrial arrhythmias but there were no differences in the proportion of patients who had hypertension, diabetes, or smoking at baseline (Table 1).

**Table 1:** Baseline Clinical Characteristics.

| *Clinical Characteristic* | *No VT/VF during*<br>*Follow Up*<br>*N=1210* | *VT/VF during*<br>*Follow Up*<br>*N=276* | *P-Value* |
| --- | --- | --- | --- |
| Age At Randomization, Mean (± SD) | 61.1 (± 12) | 59.7 (± 12) | 0.116 |
| Female Sex, N (%) | 534 (44%) | 78 (28%) | **<.001** |
| LVEF≤25, N (%) | 751 (62%) | 198 (72%) | **0.003** |
| Not Indicated for CRT | 740 (60%) | 138 (50%) | **0.002** |
| History of CHF Hospitalization, N (%) | 144 (12%) | 76 (28%) | **<.001** |
| Diabetes, N (%) | 305 (25%) | 64 (23%) | 0.468 |
| Smoking at Baseline, N (%) | 141 (12%) | 40 (15%) | 0.182 |
| History of Atrial Fibrillation, N (%) | 52 (9%) | 15 (14%) | 0.100 |
| History of NSVT, N (%) | 50 (4%) | 18 (7%) | 0.086 |
| Weight in Kilograms, Mean (± SD) | 85.1 (± 22) | 90.2 (± 24) | **0.001** |
| ACE-I or ARB, N (%) | 1124 (93%) | 263 (95%) | 0.150 |
| Beta Blocker Medication at Baseline, N (%) | 1138 (94%) | 258 (94%) | 0.720 |

Abbreviations: ACE-I: angiotensin converting enzyme-inhibitor, ARB-angiotensin receptor blocker, CHF: congestive heart failure, CRT: cardiac resynchronization therapy, LVEF-left ventricular ejection fraction, NSVT: non-sustained ventricular tachycardia, VT: ventricular tachycardia, VF: ventricular fibrillation

### Risk Score for Prediction of VTA

Table 2 shows the 4 independent predictors for increased risk of VTAs among patients with NICM. These factors were: patients with ICD not meeting indication for CRT, male sex, LVEF ≤25%, and Black race. Using the point estimates from the Fine and Gray model each variable was assigned a number of points. Male sex was assigned 6 points, ICD without CRT was assigned 3 points, LVEF≤25% was assigned 4 points, and Black race was assigned 6 points. Patients were then divided into 3 groups of risk, with low-risk patients defined as those with a score ≤4, intermediate risk as those with a score of 4-10 and high-risk patients defined as those with a score >10.

**Table 2:** Selection of Variables for Risk Score.

| *Variable* | *Hazard Ratio* | *95% Confidence interval* | *P-Value* | *Points* |
| --- | --- | --- | --- | --- |
| Male | 1.88 | 1.5 - 2.4 | <.001 | 6 |
| Not Indicated for CRT | 1.40 | 1.1 - 1.7 | 0.002 | 3 |
| Black Race | 1.61 | 1.3 - 2.1 | <0.001 | 6 |
| LVEF≤25% | 1.34 | 1.1 -1.7 | <0.001 | 4 |

CRT: cardiac resynchronization therapy
LVEF: Left ventricular ejection fraction

### Application of Risk-Score in Prediction of First Ventricular Arrhythmic Event

Figure 2A shows the cumulative incidence of Any VT/VF by each of the 3 different risk groups. There were 390, 728, and 387 patients in the low-, intermediate-, and high-risk groups respectively. The cumulative incidence of any VT/VF at 5 years was 42% in the high-risk group 24% in the intermediate risk group and 15% in the low-risk group, p<0.001 for the overall comparison. Consistently, in Fine and Gray regression analysis, the high-risk group was found to have nearly a 3-fold increased risk of Any VT/VF when compared to the low-risk group (HR=3.29, 95% CI [2.3-4.8], p<0.001). The intermediate-risk group was found to have a nearly 2-fold increased risk of Any VT/VF as the low-risk group (HR=1.90, 95% CI [2.3-4.8] p>0.001). Similar findings were observed for the end point of Fast VT/VF and appropriate ICD shock (Table 3 B, C).

**Table 3:** Prediction of First Ventricular Arrhythmic Event by Risk Score Category (Development Cohort)

| *Endpoint* | *Group Comparison* | *Hazard Ratio* | *95% Confidence Interval* | *P-Value* |
| --- | --- | --- | --- | --- |
| Any VT/VF | Intermediate vs Low | 1.90 | 1.3 -2.7 | <0.001 |
|  | High vs Low | 3.29 | 2.3 -4.8 | <0.001 |
| Fast VT/VF | Intermediate vs Low | 2.31 | 1.4 -3.7 | <0.001 |
|  | High vs Low | 3.98 | 2.5 -6.5 | <0.001 |
| Appropriate ICD Shock | Intermediate vs Low | 2.49 | 1.3 -4.8 | 0.006 |
|  | High vs Low | 4.29 | 2.2 -8.3 | <0.001 |

### Incidence of Fast VT/VF and Appropriate ICD Shocks According to Risk Score

Figure 2B shows the cumulative incidence of Fast VT/VF according to the risk score. The cumulative risk of Fast VT/VF during 5 years of follow-up was 31%, 19% and 9% in patients with a high, intermediate, and low risk score, p<0.001 for the overall comparison. Using Fine and Gray regression, intermediate-risk patients had a significant 2.31-fold increased risk of Fast VT/VF when compared to intermediate risk patients and nearly a 4-fold increased risk when compared to low-risk patients (Table 3B). Similar findings were observed for the end point of appropriate ICD shock (Figure 2C). Of note, the average yearly rates of life-threatening Fast VT/VF and appropriate ICD shock among patients with NICM in the low-risk group were only 1.8 and 0.8, respectively.

**Figure 2:**
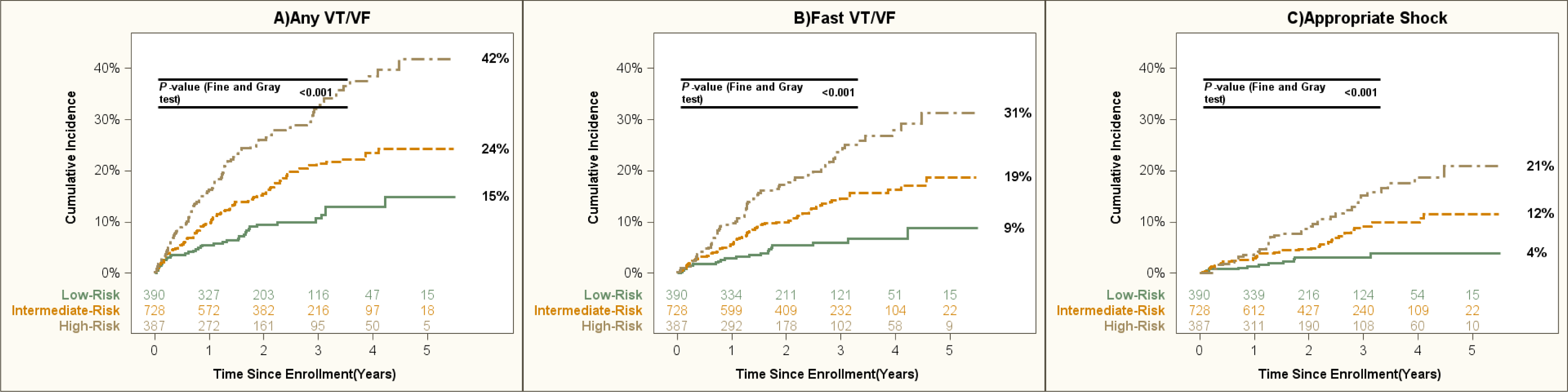
First Ventricular Arrhythmic Event in Development Cohort.

### Application of Risk-Score in Prediction of Recurrent Arrhythmic Events

A recurrent event analysis to estimate the burden of events among patients with NICM yielded consistent results. The mean number of events per patient during the 5 years of follow up was 1.5 in the high-risk group, 0.9 in the intermediate-risk group, and 0.50 in the low-risk group (Figure 3A). Consistently, the burden of events was 4.62 times higher in the high-risk group compared to the low-risk group (HR=3.39, 95% CI [2.1-5.5] P<0.001). The burden of recurrent events was more than 2-fold in the intermediate-risk group than the low-risk group (HR=2.41, 95% CI [1.4-4.1] p<0.001). Consistent results were obtained for the recurrent event analysis of any VT/VF, Fast VT/VF, and Appropriate ICD shocks (Figure 3 B, C). The mean number of appropriate ICD shocks per patient during the 5 years of follow up among those in the low-risk group was only 0.02.

**Figure 3:**
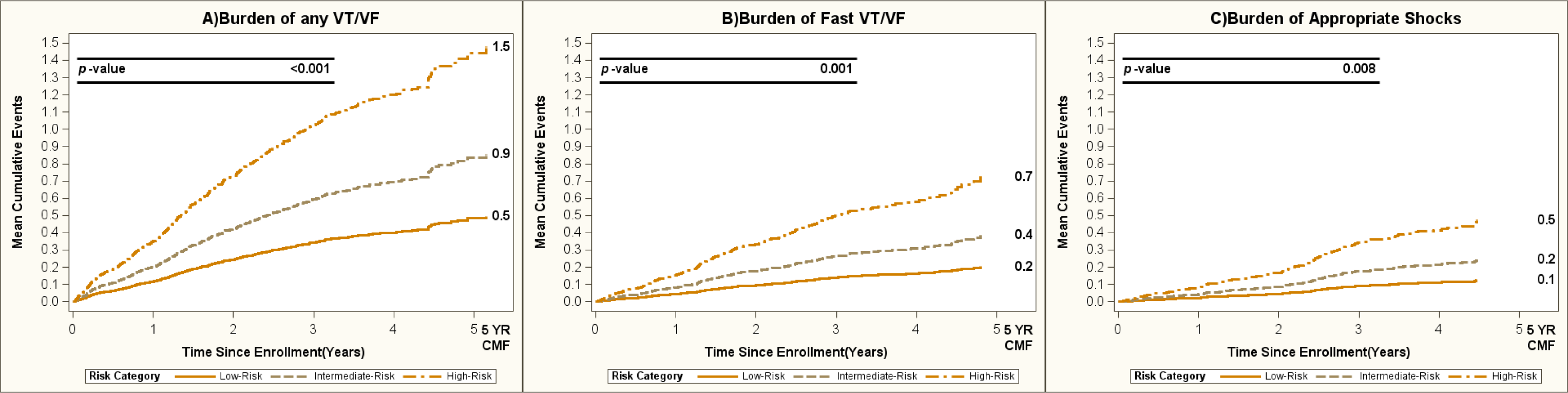
Recurrent Ventricular Arrhythmic Events in Development Cohort.

### Model validation

#### A. Internal Validation of the Model in the Development Cohort (N=1515)

Calibration plots for each of the study endpoints are shown in Supplementary Figure 1. The AUC estimations for the endpoint of any VT/VF, Fast VT/VF, and Appropriate ICD Shock were 68, 65.8, and 64.5, respectively, after 5 years of follow up (Supplementary Figure 1).

#### B. External Validation of the Model in the RAID Trial (N=332)

We ran the analysis for the validation cohort. The regression model results for the first arrhythmic event and recurrent arrhythmic events are shown in Table 5 and Table 6, respectively. The figures including the cumulative incidence function plots and the cumulative mean function for recurrent events are shown in Figure 4 and Figure 5, respectively.

**Figure 4:**
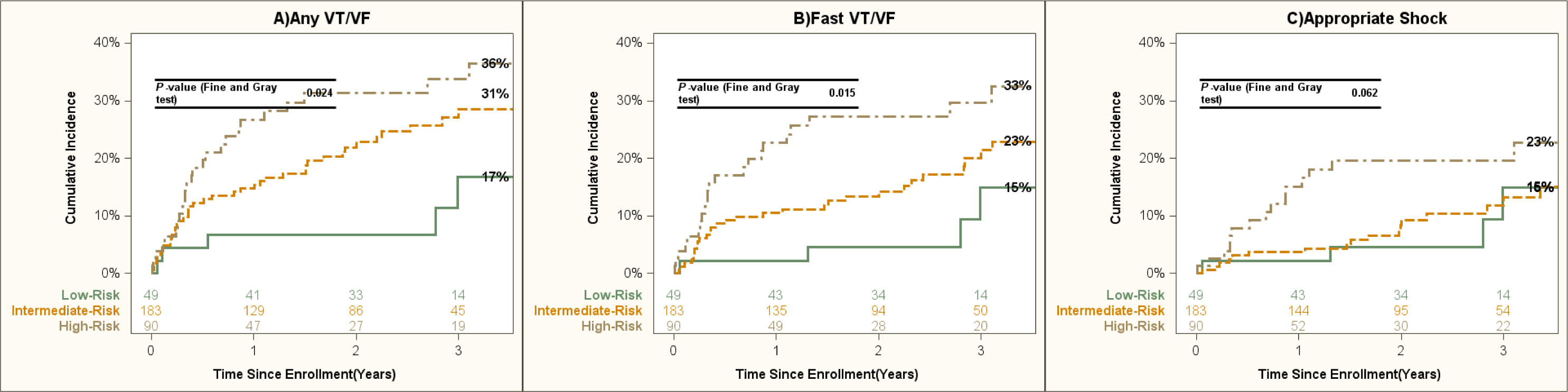
First Ventricular Arrhythmic Event in Validation Cohort.

**Figure 5:**
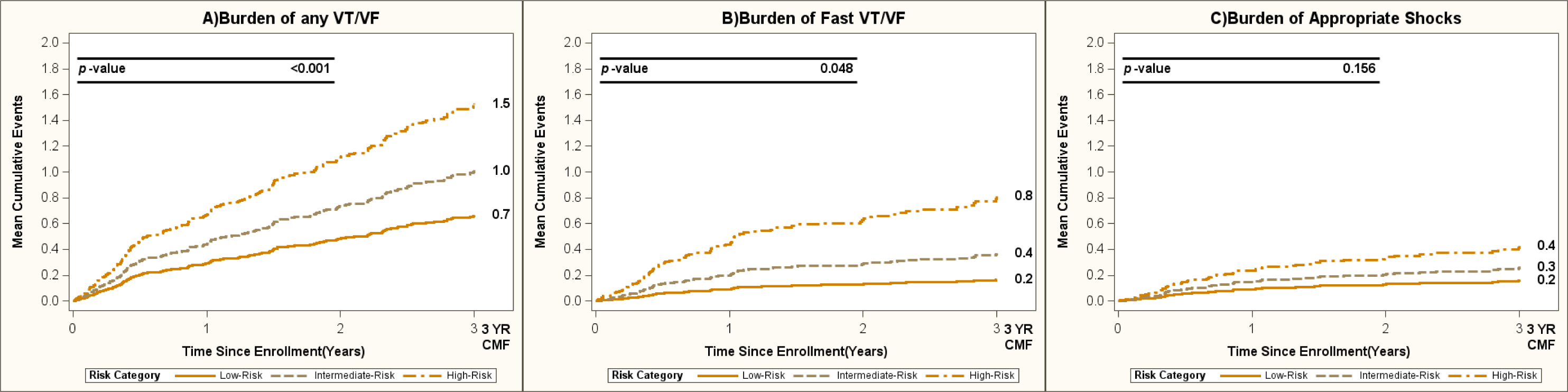
Recurrent Ventricular Arrhythmic Events in Validation Cohort.

**Table 4:** Prediction of Recurrent Ventricular Arrhythmic Events by Risk Score Category (Development Cohort)

| *Recurrent Event Endpoint* | *Group Comparison* | *Hazard Ratio* | *95% Confidence Interval* | *P-Value* |
| --- | --- | --- | --- | --- |
| Burden of Any VT/VF | Intermediate vs Low | 2.40 | 1.5 -3.9 | <0.001 |
|  | High vs Low | 3.39 | 2.1 -5.5 | <0.001 |
| Burden of Fast VT/VF | Intermediate vs Low | 2.41 | 1.4 -4.1 | 0.001 |
|  | High vs Low | 3.95 | 2.3 -6.9 | <0.001 |
| Burden of Appropriate ICD Shocks | Intermediate vs Low | 2.67 | 1.3 -5.5 | 0.008 |
|  | High vs Low | 4.40 | 2.1 -9.2 | <0.001 |

**Table 5:** RAID Validation for First Ventricular Arrhythmic Event.

| *Endpoint* | *Group Comparison* | *Hazard Ratio* | *95% Confidence Interval* | *P-Value* |
| --- | --- | --- | --- | --- |
| Any VT/VF | Intermediate vs Low | 2.46 | 1.0 -6.2 | 0.055 |
|  | High vs Low | 3.48 | 1.3 -9.1 | 0.011 |
| Fast VT/VF | Intermediate vs Low | 2.11 | 0.8 -5.9 | 0.152 |
|  | High vs Low | 3.80 | 1.3 -10.9 | 0.013 |
| Appropriate ICD Shock | Intermediate vs Low | 1.21 | 0.4 -3.5 | 0.732 |
|  | High vs Low | 2.55 | 0.8 -7.7 | 0.095 |

**Table 6:** Recurrent Ventricular Arrhythmic Events (Validation Cohort)

| *Recurrent Event Endpoint* | *Group Comparison* | *Hazard Ratio* | *95% Confidence Interval* | *P-Value* |
| --- | --- | --- | --- | --- |
| Burden of any VT/VF | Intermediate-Risk vs Low-Risk | 4.82 | 1.9 -11.9 | <0.001 |
|  | High-Risk vs Low-Risk | 4.62 | 1.8 -12.0 | 0.002 |
| Burden of Fast VT/VF | Intermediate-Risk vs Low-Risk | 3.06 | 1.0 -9.3 | 0.048 |
|  | High-Risk vs Low-Risk | 6.02 | 1.9 -19.1 | 0.002 |
| Burden of Appropriate ICD Shocks | Intermediate-Risk vs Low-Risk | 2.26 | 0.7 -7.0 | 0.156 |
|  | High-Risk vs Low-Risk | 3.09 | 1.0 -9.7 | 0.053 |

Calibration plots for each of the study endpoints are shown in Supplementary Figure 2. The AUC estimations for the endpoint of any VT/VF, Fast VT/VF, and Appropriate ICD Shock were 66.7, 67.7, and 69.3 after 2.8 years of follow up.

## DISCUSSION

In this study, which included 1515 patients with NICM from three major ICD trials, we developed a risk stratification score for the occurrence of a first VT/VF, based on four simple and readily available demographic and clinical risk factors, that is specific for patients with NICM. The applicability of the model was assessed for the more severe endpoints of Fast VT/VF (defined as VT ≥ 200 bpm or VF) and appropriate ICD shocks and was externally validated in the contemporary RAID population. We have identified 3 main risk groups for the occurrence of any VT/VF, fast VT/VF, and appropriate ICD shock in NICM patients who received a primary prevention ICD, based on 4 simple factors: No-CRT indication, male sex, LVEF ≤25%, and Black race). Our findings have several important clinical implications. First, patients with NICM still have an overall pronounced risk of VT/VF during follow up, approaching 27% at 5 years (average yearly rate of approximately 5.4%). Second, patients with NICM can be stratified into high, intermediate, and low risk strata based on the presence of 4 clinical variables including LVEF, race, ICD implantation and sex. Finally, it should also be noted that in the low risk group the average yearly rate of more severe life-threatening arrhythmic events, including Fast VT (≥200 bpm) or VF and appropriate ICD shocks, were only1.8 and 0.8, respectively. These data can be used for informed decision making when discussing the need for primary ICD implantation in device candidates with NICM.

The DANISH trial called into question the benefit of an ICD in patients with NICM as it showed that there was no significant difference in the overall risk of death between ICD and non-ICD recipients(3). It should be noted that the DANISH trial did not account for competing risks, and 31% of deaths in the trial were due to non-cardiovascular causes. In our study we specifically evaluated the risk of ventricular tachyarrhythmia in patients with NICM while accounting for the competing risk of death without prior VT/VF. Moreover, using available clinical and echocardiographic variables we were able to risk stratify non-ischemic patients into high, intermediate, and low risk groups. Nevertheless, using the risk factors identified in our study we were able to identify a group of patients at relatively low risk for appropriate ICD shocks (4% at 5 years). Of note, in MADIT-RIT only therapy for Fast VT/VF and appropriate ICD shocks were found to be lifesaving, whereas ICD therapy for lower rate VT/VF events was associated with a significant increase in the risk of death (6). Thus, based on our findings, it is expected that most patients with NICM in the low-risk group will not utilize the ICD for a lifesaving therapy during long-term follow-up.

Male sex has also been associated with an increased risk of VT/VF in our score. Evaluating sex differences in the risk of VT/VF among patients with NICM is challenging as there is a paucity of data in women(11). However, in landmark trials evaluating ICD primary prevention therapy, women comprised a higher proportion of patients with NICM than patients with ICM(11,12). Contemporary data suggest that women with NICM treated with ICD have a lower cumulative incidence of VT/VF compared with men(11,13). This discrepancy in the susceptibility to VT/VF between women and men may be secondary to sex differences in ventricular repolarization, autonomic tone, ion channel function, intracellular calcium handling, and excitation contraction coupling(11,14).

Our study identified Black patients as having increased arrhythmic risk. This finding maybe related to other social determinants of health that were not captured in the trial database. In a sub-analysis of the Sudden Cardiac Death-Heart Failure Trial (SCD-HeFT), there was no difference by race in the proportion of ICD shocks (15). As with women, Black patients have been underrepresented in most major large randomized heart failure clinical trials(15,16), therefore the power to detect a difference is greatly attenuated. In a prospective cohort of patients undergoing ICD implantation for primary prevention of SCD, Black patients had an increased risk of dying without receiving an appropriate ICD shock compared to non-Black patients(17). Our results suggest that, while accounting for competing risks, the risk of VTAs is greatly increased in Black patients and therefore is a significant component of our score. These data support the use of prophylactic ICD placement in black patients with NICM.

Finally, LVEF ≤ 25% was another factor in the scoring system predicting VT/VF. An analysis of the Prolongation of Reverse Remodeling Period to Avoid Untimely ICD implantation in Newly Diagnosed Heart Failure Using the Wearable Cardioverter Defibrillator (PROLONG) study showed that patients with newly diagnosed NICM and severely reduced EF experienced an elevated risk for VT/VF during the early phase of optimization of HF therapy(18). Despite this elevated risk, early device implantation does not seem to improve survival in patients with NICM(18,19). The reason may be attributed to the potential for LVEF recovery in patients with newly diagnosed NICM(18,19).This phenomenon highlights the heterogeneity that is inherently characteristic of the patient population with NICM(20). Patients with severely reduced LVEF after the early risk stratification period may represent a subgroup of NICM with persistent myocardial damage that increases their risk for subsequent VTAs.

This study has important clinical implications that pertain both to guiding decision making when evaluating patients with NICM for a primary prevention ICD and risk stratification during follow up. Given the controversial trial data that exist to date regarding the benefit of an ICD in patients with NICM, our study provides clinicians the ability to risk stratify such patients into low-intermediate and high-risk groups. The clinical risk-score that was constructed adds objective data from major multicenter, randomized ICD trials which may be incorporated into the shared decision-making process and may help patients proceed with a decision in greater confidence. We have additionally shown that the patients in the high and intermediate-risk groups not only experience a higher incidence of VTAs but also a greater burden of events when compared to the low-risk group (Central Illustrations). These results suggest that even after a decision for ICD implantation has been made, the risk score can be used to determine which patients may need to be followed more closely.

### Limitations

Our study has several limitations that require acknowledgement. First, the analysis was performed in a post hoc fashion after combining patient data from several different trials with patient enrollment during different time periods. The risk of SCD in HFrEF continues to decline with evolving guideline-directed medical therapy for HF(10,21,22). Therefore, the results of this study are hypothesis generating and warrant further investigation in the form of a prospective interventional study in contemporary settings. That being said, it must be noted that data from the National Cardiovascular Registry showed that only 61% of patients filled any GDMT prior to implant(23). Additionally, most patients are not being treated with the appropriate target doses of GDMT for HFrEF(24), and this suggests that a contribution of medical therapies in a real-world setting to the reduction in SCD may be attenuated due to prescription, compliance, and adherence with appropriate treatment, thereby, not obviating the need for a primary prevention ICD. Nevertheless, our prediction model was externally validated in the more contemporary RAID population (published in 2018), thereby providing greater confidence in the applicability of our findings to contemporary patients with NICM. Second, improvements in LVEF with cardiac resynchronization can reduce the risk of ventricular tachyarrhythmia (25). We did not have routine follow-up LVEF data to assess the impact of subsequent left ventricular function as a time dependent variable in our prediction model. Finally, there is recent evidence of cardiovascular magnetic resonance imaging (CMRI) can facilitate the identification of ventricular arrhythmogenic substrates in NICM patients(26). Unfortunately, such imaging data were not collected as part of the MADIT trials. Nevertheless, we offer four simple clinical factors that can be used for evaluating ICD candidacy without the need for more advanced and costly testing.

### Conclusions and Clinical Implications

Our findings suggest that patients with NICM who are ICD candidates experience a significant risk for VT/VF and a high burden of VT/VF during follow-up. However, simple demographic and clinical variables can be used to risk stratify patients for the occurrence of VT/VF. We identified a relatively low-risk group of patients with NICM who experience a low rate of appropriate ICD shocks during long-term follow-up. Current guidelines stress the importance of patient-physician shared decision making among candidates for a primary prevention ICD. We believe that our findings can be used to supplement shared decision making in patients with NICM who are considered for prophylactic ICD placement.

#### Perspectives-Clinical Competencies

##### Competency in Medical Knowledge

Patients with non-ischemic cardiomyopathy who are implanted with an ICD for primary prevention of sudden cardiac death experience both an increased risk for first and recurrent ventricular arrhythmic events.

##### Competency in Patient Care

Simple clinical variables can be used to predict the ventricular arrhythmic risk of patients with non-ischemic cardiomyopathy and can help support patient-physician shared decision making with regard to prophylactic ICD placement.

##### Translational Outlook

Treatment of heart failure and arrhythmias has evolved significantly over the last 20 years and a prospective randomized trial is warranted to evaluate whether ICD is effective in reducing sudden cardiac death in contemporary patients with non-ischemic cardiomyopathy who are receiving current guideline-directed medical therapy.

## Data Availability

The original contributions presented in this study are included in the article/Supplementary material, further inquiries can be directed to the corresponding author.

## Acknowledgments

Ido Goldenberg: None, Arwa Younis: None, David Huang: Research grants from Medtronic, Biosense-Webster, and Biotronik. Spencer Z. Rosero: Research grants from Medtronic and Biotronik. Valentina Kutyifa: Research grants from Boston Scientific, Biotronik, and ZOLL. Scott McNitt: None. Bronislava Polonsky: None. Jonathan S. Steinberg: Research grants from NIH, Medtronic, AliveCor and Atricure. Wojciech Zareba: Research grants from Boston Scientific. Ilan Goldenberg: Research grants from Boston Scientific, Zoll, Medtronic, Biosense-Webster, and Biotronik . Mehmet K. Aktas: Research grants from Boston Scientific and Medtronic

## Abbreviations

HF: Heart Failure
ICD: implantable cardioverter defibrillator
NICM: non-ischemic cardiomyopathy
VTA: ventricular tachyarrhythmia
MADIT: multicenter automated defibrillator trial
RAID: ranolazine in high-risk patients with implantable cardioverter defibrillators
CRT: D-cardiac resynchronization therapy with a defibrillator

---

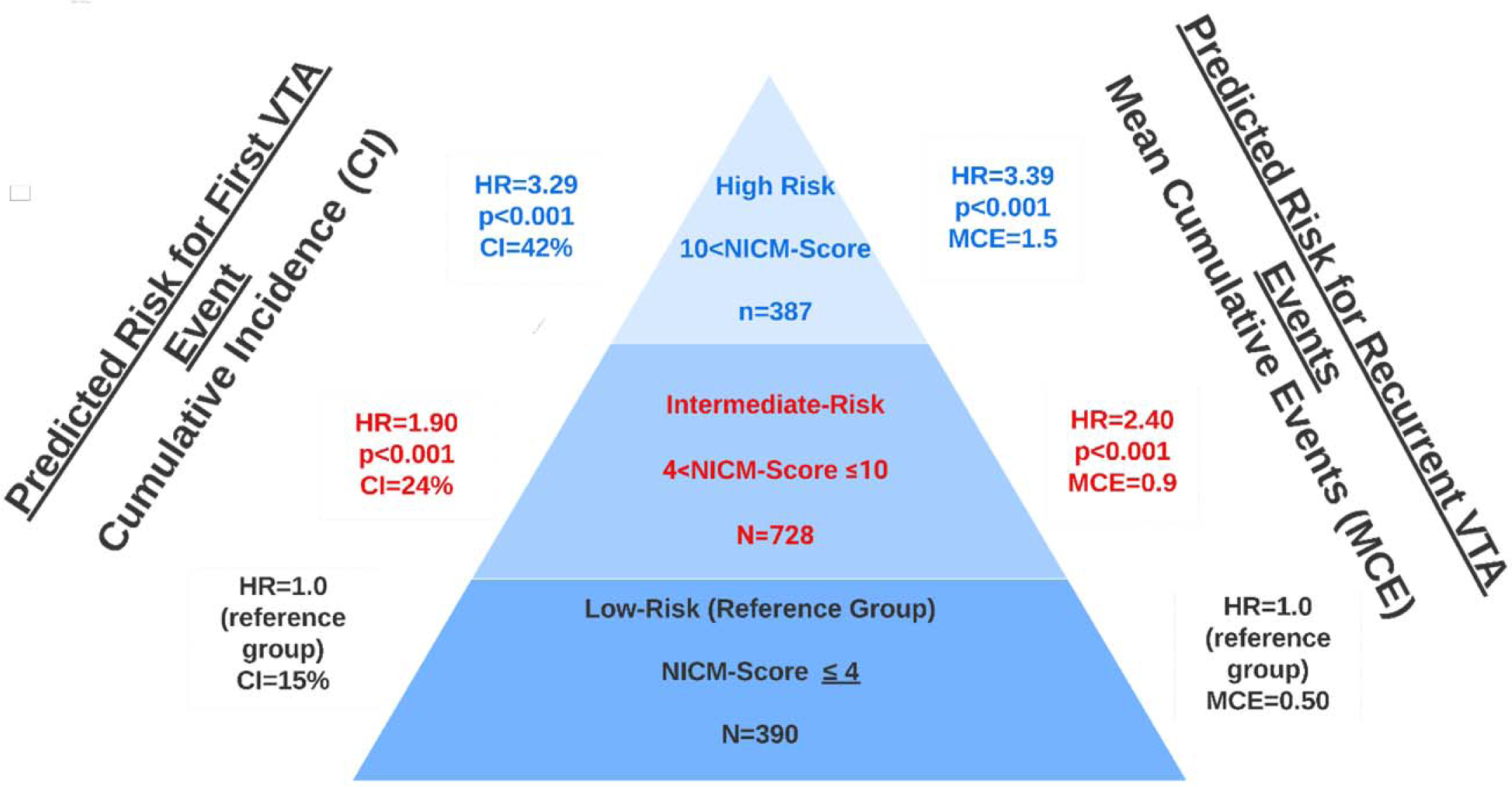
Central Illustration: NICM-Score Predicts Risk of VTA (Development Cohort)

Supplementary Figure 1: Calibration Plots and Internal Validation for Development Cohort

Supplementary Figure 2: Calibration Plots and External Validation for Validation Cohort

